# Four years of PrEP use; sexual behaviour and STIs in the AMPrEP demonstration project cohort among men who have sex with men in Amsterdam, the Netherlands

**DOI:** 10.1101/2023.12.11.23299798

**Authors:** Mark A.M. Van Den Elshout, Eline Wijstma, Anders Boyd, Vita Jongen, Liza Coyer, Peter L. Anderson, Udi Davidovich, Henry J.C. De Vries, Maria Prins, Maarten F. Schim Van Der Loeff, Elske Hoornenborg, the Amsterdam PrEP Project team in the HIV Transmission Elimination AMsterdam Initiative (H-TEAM)

**Affiliations:** Department of Infectious Diseases, Public Health Service of Amsterdam, Amsterdam, the Netherlands; Stichting hiv monitoring, Amsterdam, the Netherlands; Department of Pharmaceutical Sciences, University of Colorado Anschutz Medical Campus, Aurora, CO, USA; Department of Social Psychology, University of Amsterdam, Amsterdam, the Netherlands; Amsterdam UMC location University of Amsterdam, Amsterdam institute for Infection & Immunity (AII), Meibergdreef 9, Amsterdam, the Netherlands; Amsterdam UMC location University of Amsterdam, Department of Dermatology, Meibergdreef 9, Amsterdam, the Netherlands; Amsterdam UMC location University of Amsterdam, Department of Infectious Diseases, Meibergdreef 9, Amsterdam, the Netherlands

**Keywords:** HIV, pre-exposure prophylaxis, PrEP, Men Who Have Sex With Men, Sexual behaviour, sexually transmitted infections, hepatitis C, sexual empowerment

## Abstract

**Background:** An increasing number of countries are currently implementing or scaling-up HIV pre-exposure prophylaxis (PrEP) care. With the introduction of PrEP, there was apprehension about possible risk compensation, particularly on the long term. To inform sexual health counselling and STI screening programmes, we aimed to study sexual behaviour and STI incidence among men who have sex with men (MSM) and transgender women who use long-term daily or event-driven PrEP.

**Methods:** The Amsterdam PrEP demonstration project (AMPrEP) provided oral daily PrEP and event-driven PrEP to MSM and transgender women in 2015-2020. Participants could choose their PrEP regimen, and could switch at each three-monthly visit. STI testing occurred at and, upon request, in-between 3-monthly study visits. We assessed changes in number of sex partners and condomless anal sex acts over time with casual partners (CAS) using negative binomial regression. We assessed changes in incidence rates (IR) of any STI (i.e., chlamydia, gonorrhoea, or infectious syphilis), individual STIs, and HIV over time using Poisson regression.

**Findings:** 367 participants (365 MSM) commenced PrEP and were followed for a median 3.9 years (IQR=3.4-4.0). The number of sex partners decreased over time (adjusted rate ratio [aRR]=0.89/year, 95%CI=0.86-0.92), while the number of CAS acts with casual partners remained stable (aRR=0.98/year, 95%CI=0.94-1.01). IR of any STI was 87/100 person years (PY) (95%CI=82-93) and did not change over time for both daily PrEP or event-driven PrEP users. However, IRs of chlamydia and gonorrhoea decreased slightly in daily PrEP users. Two daily PrEP users, and no event-driven PrEP users, were diagnosed with HIV during their first year on PrEP.

**Conclusion:** With no increase in the number of casual sex partners nor of CAS acts, we found no indication of possible risk compensation during the first four years of PrEP use. Although the STI incidence was high, it did not increase over time.

**Funding:** ZonMw, RIVM, GGD, H-TEAM, Gilead.

## Introduction

Since the publication of results from the iPrEX study in 2010(1), data on oral pre-exposure prophylaxis (PrEP) use to prevent HIV have been accumulating through various randomised clinical trials (RCTs), demonstration studies and implementation projects. PrEP has shown to be acceptable, safe and highly effective in preventing HIV acquisition, provided adherence is good, especially among men who have sex with men (MSM)(2, 3). As such, PrEP plays a pivotal role in achieving the UNAIDS goal of zero new HIV infections(4, 5), and is being rolled out in an increasing number of countries(6).

With the introduction of PrEP there was apprehension about possible risk compensation. In 2018, a systematic review of 17 PrEP cohort studies among MSM reported increases in condomless sex among PrEP users and increased sexually transmitted infections (STI) diagnoses, but the median follow-up time of included studies was only 6 months (range 3-18 months)(7). One of the larger studies to date reported stable STI incidence, despite increased receptive condomless sex acts, with a median follow-up of 22 months(8). To the best of our knowledge, there are no studies with longer-follow-up time evaluating behavioural trends and STI incidence rates among MSM on PrEP. However, since many PrEP users are expected to use PrEP for several years, such information would be needed to inform policy makers and clinicians of current and future PrEP programmes. Therefore, we prospectively assessed sexual behaviour and incidence rates of HIV and other STIs, including hepatitis C virus (HCV), among MSM on PrEP for up to four years. We also assessed switching between the daily and event-driven regimen, PrEP discontinuation and adherence to PrEP.

## Methods

### Study design

The Amsterdam PrEP demonstration project (AMPrEP) was an open-label demonstration study conducted between 3 August 2015 and 1 December 2020 that included MSM and transgender women. Participants were offered a free-of-charge oral coformulation of emtricitabine and tenofovir disoproxil 200/245 mg (TDF/FTC) to be used as daily or event-driven PrEP. The study design, aim and procedures have been described previously(9), and analyses of the first 24 months of follow-up and HCV incidence have been published before(10, 11). Briefly, participants attended 3-monthly study visits at the Centre for Sexual Health of the Public Health Service of Amsterdam, the Netherlands.

Eligible were MSM and transgender women without HIV, who were ≥18 years old and had, in the 6 months prior to screening, a substantial likelihood to acquire HIV sexually(9). Switching between daily PrEP and event-driven PrEP was allowed at each 3-monthly study visit. All AMPrEP participants provided samples for HIV, HCV and STI testing at each study visit. We requested participants to also provide blood for dried blood spots (DBS) to measure adherence at the 3 or 6 and 12, 24 and 48 months study visits. We tested for HCV every 12 months until December 2016, and bi-annually thereafter(11, 12).

### Measures

Sociodemographic, psychosocial, clinical and behavioural characteristics were collected via questionnaires. Demographics collected at inclusion in AMPrEP were age, gender identity, self-declared ethnicity, place of residency, education level, employment status, income level, living situation, relationship status and sexual preference. Behavioural and clinical characteristics included history of condomless anal sex and bacterial STIs in the 6 months prior to inclusion. Self-reported number of sex partners and anal sex acts, including partner type and condom use, were recorded three-monthly. Participants self-reported half-yearly whether they engaged in chemsex, defined as the use of γ-hydroxybutyrate/γ-butyrolactone, methamphetamine or mephedrone prior to or during sex.

Psychosocial determinants were measured yearly. Sexual compulsivity was measured using the sexual compulsivity scale(13), with a score ≥24 being indicative of a greater impact of sexual thoughts on daily functioning and of an inability to control sexual thoughts or behaviours(14). Sexual satisfaction was measured using the New Sexual Satisfaction Scale (NSSS) on a discrete scale(15).

Symptoms of depression or anxiety were assessed using the Mental Health Inventory-5 (MHI-5) score, where a score of <60 indicated symptoms of depression or anxiety(16). The Alcohol Use Disorders Identification Test (AUDIT)(17) and Drug Use Disorder Identification Test (DUDIT)(18) questionnaires were used to assess problematic alcohol and drug use, respectively; scores ≥8 are interpreted as indicative of alcohol-related or drug-related problems(19).

### Outcomes

We assessed the number of sex partners, number of anal sex acts, and number of condomless anal sex (CAS) acts with casual partners in the past 3 months at each study visit.

We assessed the number of diagnoses of chlamydia, gonorrhoea, infectious syphilis (stage 1, 2 and recent latent syphilis), HCV and HIV. We calculated incidence rates as the number of visits with a diagnosis (including repeat infections) divided by the person years (PY) of follow-up. Diagnoses were laboratory-confirmed infections from samples taken during study visits or additional visits at the Centre for Sexual Health during follow-up. We defined any STI as having one or more bacterial STIs (i.e., chlamydia, gonorrhoea or infectious syphilis) at a visit. We stratified chlamydia and gonorrhoea infections by anatomical site (i.e., anal, urogenital, or pharyngeal), and defined any anal STI as having anal chlamydia or anal gonorrhoea. In calculating PY for IRs of bacterial STI, we assumed that infection occurred at the date of positive test and follow-up time recontinued after infection. We defined incident HCV infections according to clinical practice guidelines(20), and distinguished between primary infections and reinfections. In calculating PY for IRs of HCV, we assumed that the infection occurred midway between the last negative and first positive test. Follow-up time stopped after infection and continued after confirmed sustained virologic response. In calculating PY for IRs of HIV, we assumed that infection occurred midway between the last negative and first positive test, and follow-up time stopped after infection.

We evaluated the number and rates of any switch between regimens as well as switch from daily PrEP to event-driven PrEP and *vice versa*. We also evaluated the number and rate of PrEP discontinuations, which were defined as one of the following: (a) a duration between study visits lasting at least nine months without self-reporting continuing PrEP elsewhere during this period, (b) reporting not having taken PrEP for at least 3 months (regardless of visit attendance), (c) attending a formal study exit visit without self-reporting continuation of PrEP elsewhere, or (d) being lost-to-follow up. Loss-to-follow-up was defined as not attending a study visit in the nine months prior to 15 March 2020 (i.e., the start of the COVID-19 lockdown measures in the Netherlands), whilst not having completed the 48-month visit. Finally, we calculated the proportion of participants who still used PrEP at 48 months after enrolment among those who could have reached 48 months of follow-up before censoring.

We calculated median levels of intracellular tenofovir diphosphate (TFV-DP) in dried blood spots and corresponding IQRs among daily PrEP users and report the proportion of daily PrEP users with good adherence (TFV-DP ≥700 fmol/punch)(21). We do not report these outcomes for event-driven PrEP users, since TFV-DP does not indicate prevention-effective adherence to event-driven PrEP(22).

### Laboratory methods

Laboratory methods were described previously(10, 23). For DBS analyses of intracellular TFV-DP, the 48 month samples were measured using a 50:50 methanol:water extraction. Results were divided by 1.138 in order to compare them with the previous 70:30 extractions.

### Statistical methods

We excluded participants without any follow-up study visits. Follow-up began at PrEP initiation (i.e., ‘baseline’) and continued until 48-months of individual follow-up, last study visit, HIV diagnosis, or 15 March 2020, whichever occurred first. We excluded periods between PrEP discontinuation (as described above) and PrEP re-initiation from follow-up time. We presented analysis for both the overall study population and stratified on daily PrEP or event-driven PrEP. PrEP regimen was included as a time-updated variable.

To analyse changes in sexual behaviour, we report the median and interquartile ranges (IQR) of sexual behaviour outcomes for each study visit. We modelled the year-on-year change in mean sexual behaviour outcomes (excluding baseline visits) using a mixed-effects negative binomial regression model with a random intercept and random slope on the participant-level to account for between-individual variability at baseline and during follow-up, respectively, and robust standard errors to ensure variance corresponded to individuals. We report relative ratios (RR) and corresponding 95% confidence intervals (CI), and used a Wald χ^2^ test to test for changes over time. We provide unadjusted estimates and estimates adjusted for age category at baseline (i.e., ≤34, 35– 44, ≥45 years).

We calculated the incidence rate difference between daily PrEP and event-driven PrEP users and report two-sided p-values. To analyse changes in bacterial STI incidence, we calculated STI IRs per 100PY of follow-up and corresponding 95%CI based on a Poisson distribution. We modelled the change in STI IRs compared to the first year on PrEP using Poisson regression with a gamma-distributed frailty, and added a random intercept on the individual level. We report incidence rate ratios (IRR) and corresponding 95%CI per year, and used a Wald χ^2^ test to test for changes compared to the first year on PrEP. We provide unadjusted estimates and estimates adjusted for age category at baseline and STI testing frequency (i.e., ≤3, 4–5, ≥6 tests per year) as a time-updated variable. We did not model change in HIV and HCV incidence rates over time owing to the low number of infections.

To analyse changes in PrEP use, we calculated regimen switch rates per 100PY and 95%CI based on a Poisson distribution, and calculated linear change in switch rates per year as switch rate ratio. We calculated the total number of PrEP discontinuations (including discontinuations after re-initiating) and median time until first discontinuing PrEP. We evaluated factors associated with time until first stopping PrEP using multivariable Cox regression, which were selected *a priori*(*24, 25*) and included the following: demographic (age, education level and place of residence), sexual behaviour and STI (number of CAS acts with casual partners, any bacterial STI in the past 3 months), and mental wellbeing. We included education level and place of residency as time-fixed variables and all other variables as time-updated. Data from (half-)yearly questionnaires were carried backwards for study visits that occurred in the past 6 and 12 months, respectively.

In sensitivity analyses, we re-ran the models on STI incidence and sexual behaviour using follow-up time including periods between PrEP discontinuation and re-initiation as periods with missing data.

We defined significance at a p-value <0.05. All statistical analyses were performed in STATA version 17.0 (StataCorp, College Station, Texas, USA). The study was registered at the Netherlands Trial Register (NL5413).

## Results

### Participants and baseline characteristics

Between August 3, 2015 and May 31, 2016, 376 participants were enrolled. Of these, nine (2.4%) were excluded from analyses because they did not attend any follow-up visits. Of the 367 included participants, 365 were MSM and two identified as transgender women. Median age at baseline was 40 years (IQR=32-48), 315/367 (86%) self-declared to be white and 280/367 (76%) had at least a college degree (S1 Table). The median follow-up time was 3.9 years (IQR=3.4-4.0), totalling 1258PY of observation. Of 282 participants who could have reached 48 months of follow-up before censoring, 192 (68%) still used PrEP after 48 months.

### Sexual behaviour

Reported numbers of sex partners and anal sex acts decreased over time (adjusted rate ratio [aRR] 0.89/year [95% CI 0.86-0.91] and 0.91/year [95% CI 0.89-0.94], respectively; Table 1). These changes were also statistically significant when stratified by PrEP regimen (Table 1, Fig 1). Number of CAS acts with casual partners did not significantly change over time (aRR 0.98/year [95% CI 0.94-1.01]), nor when stratified by regimen (Table 1, Fig 1). Numbers of sex partners and CAS acts with casual partners were higher in daily PrEP users compared to event-driven PrEP users (S2 Table).

**Fig 1.**
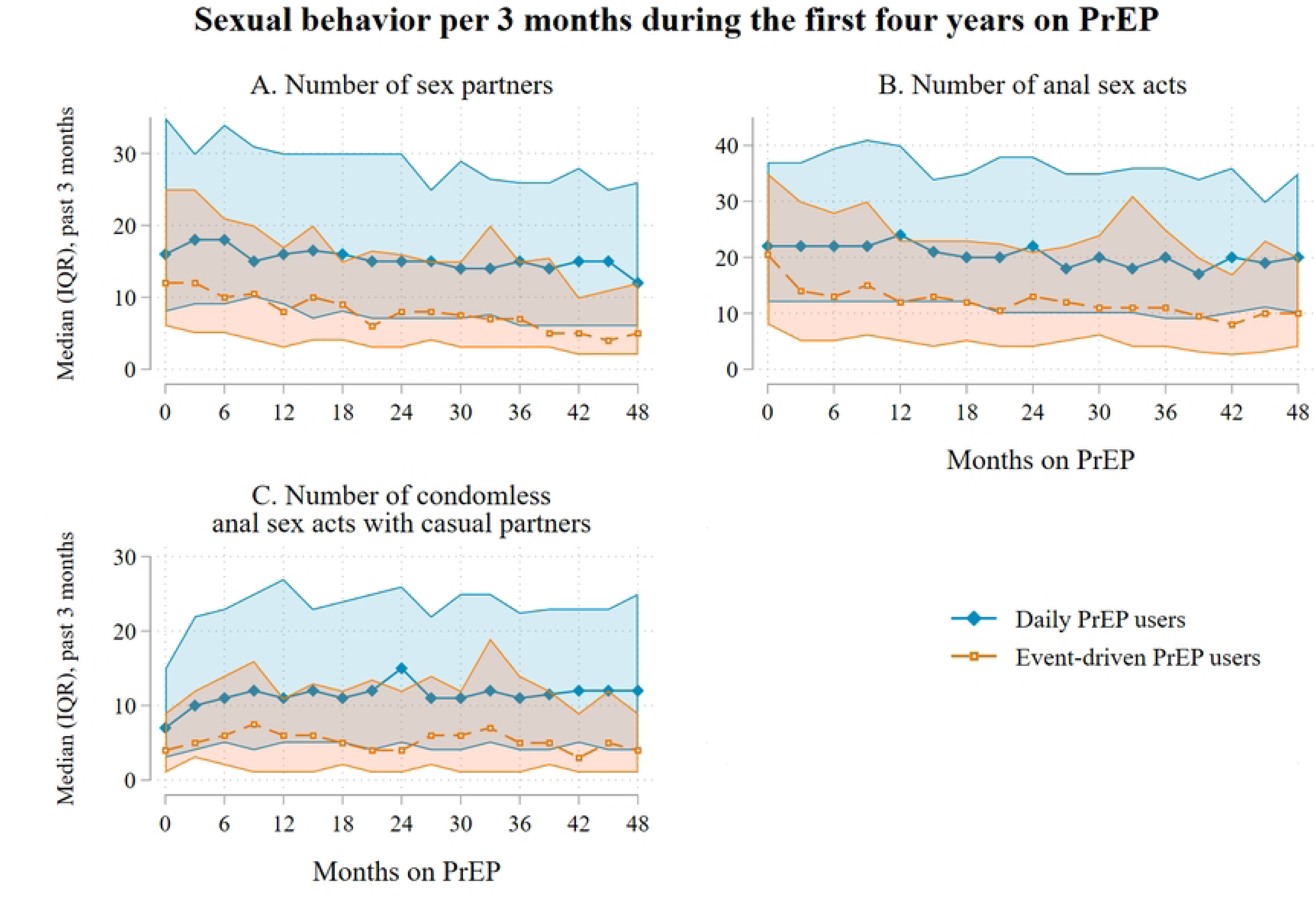
Sexual behaviour during the first four years on PrEP, AMPrEP, The Netherlands, 2015-20. Panel A shows median (IQR) number of sex partners, panel B shows median number of anal sex acts, and panel C shows median number of CAS acts with casual partners. Lines represent medians per study visit. Shaded regions represent interquartile ranges. Abbreviations: AMPrEP: Amsterdam PrEP demonstration project; IQR: interquartile range; PrEP: pre-exposure prophylaxis.

**Table 1.** Four-year outcomes of sexual behaviour per year on PrEP among AMrEP participants, Amsterdam, e Netherlands, 2015-20.

|  |  | Crude |  |  | Adjusted<sup>a</sup> |  |  |
| --- | --- | --- | --- | --- | --- | --- | --- |
|  | No. of visits with data | Crude RR (95% CI) |  | p-value | aRR (95% CI) |  | p-value |
| Number of sexual partners<sup>b</sup> |  |  |  |  |  |  |  |
| Overall | 5,105 | 0.89 | [0.86-0.91] | <0.0001 | 0.86 | [0.83-0.88] | <0.0001 |
| Daily | 3,787 | 0.91 | [0.88-0.94] | <0.0001 | 0.87 | [0.84-0.91] | <0.0001 |
| Event-driven | 1,318 | 0.85 | [0.81-0.90] | <0.0001 | 0.82 | [0.77-0.88] | <0.0001 |
| Number of anal sex acts<sup>b</sup> |  |  |  |  |  |  |  |
| Overall | 5,112 | 0.91 | [0.88-0.93] | <0.0001 | 0.88 | [0.85-0.91] | <0.0001 |
| Daily | 3,791 | 0.93 | [0.90-0.96] | <0.0001 | 0.90 | [0.87-0.93] | <0.0001 |
| Event-driven | 1,321 | 0.85 | [0.80-0.90] | <0.0001 | 0.84 | [0.79-0.89] | <0.0001 |
| Number of CAS acts with casual partners<sup>b</sup> |  |  |  |  |  |  |  |
| Overall | 5,108 | 0.97 | [0.94-1.01] | 0.17 | 0.92 | [0.88-0.97] | 0.0006 |
| Daily | 3,787 | 1.00 | [0.96-1.04] | 0.96 | 0.94 | [0.89-0.98] | 0.018 |
| Event-driven | 1,321 | 0.93 | [0.87-1.00] | 0.067 | 0.90 | [0.82-0.98] | 0.012 |

Abbreviations: AMPrEP: Amsterdam PrEP demonstration project; CAS: condomless anal sex; CI: confidence
interval; PrEP: pre-exposure prophylaxis; (a)RR: (adjusted) rate ratio.
<sup>a</sup>Adjusted for age category at enrolment (i.e., <35, 35-44, ≥45 years), and including a random intercept and random slope at the participant-level.
<sup>b</sup>In the past 3 months

Results from sensitivity analyses assessing behaviour over time since initiating PrEP while including periods without PrEP use or follow-up were largely the same, and additionally showed a decrease in the number of CAS acts with casual partners among event-driven users (aRR 0.92 [95%CI 0.85-0.99]; p=0.032; S3 Table).

### Incidence of bacterial sexually transmitted infections

In 1092 consultations among 289/367 participants, at least one STIs was diagnosed during 1258PY: 891 during 914PY among daily PrEP users and 201 during 344PY among event-driven PrEP users. IR of any STI was 87/100PY (95%CI=82-92), 97/100PY for daily PrEP users (95%CI=91-104) and 59/100PY for event-driven PrEP users (95%CI=51-67; Fig 2; Table 2; S4 Table). Compared to the first year, IR of any STI was lower in the second (aIRR=0.83, 95%CI=0.71-0.98) and third (aIRR=0.83, 95%CI=0.70-0.98) year (S5 Table). This decrease was not seen in the fourth year (aIRR=0.95, 95%CI=0.79-1.10).

**Fig 2.**
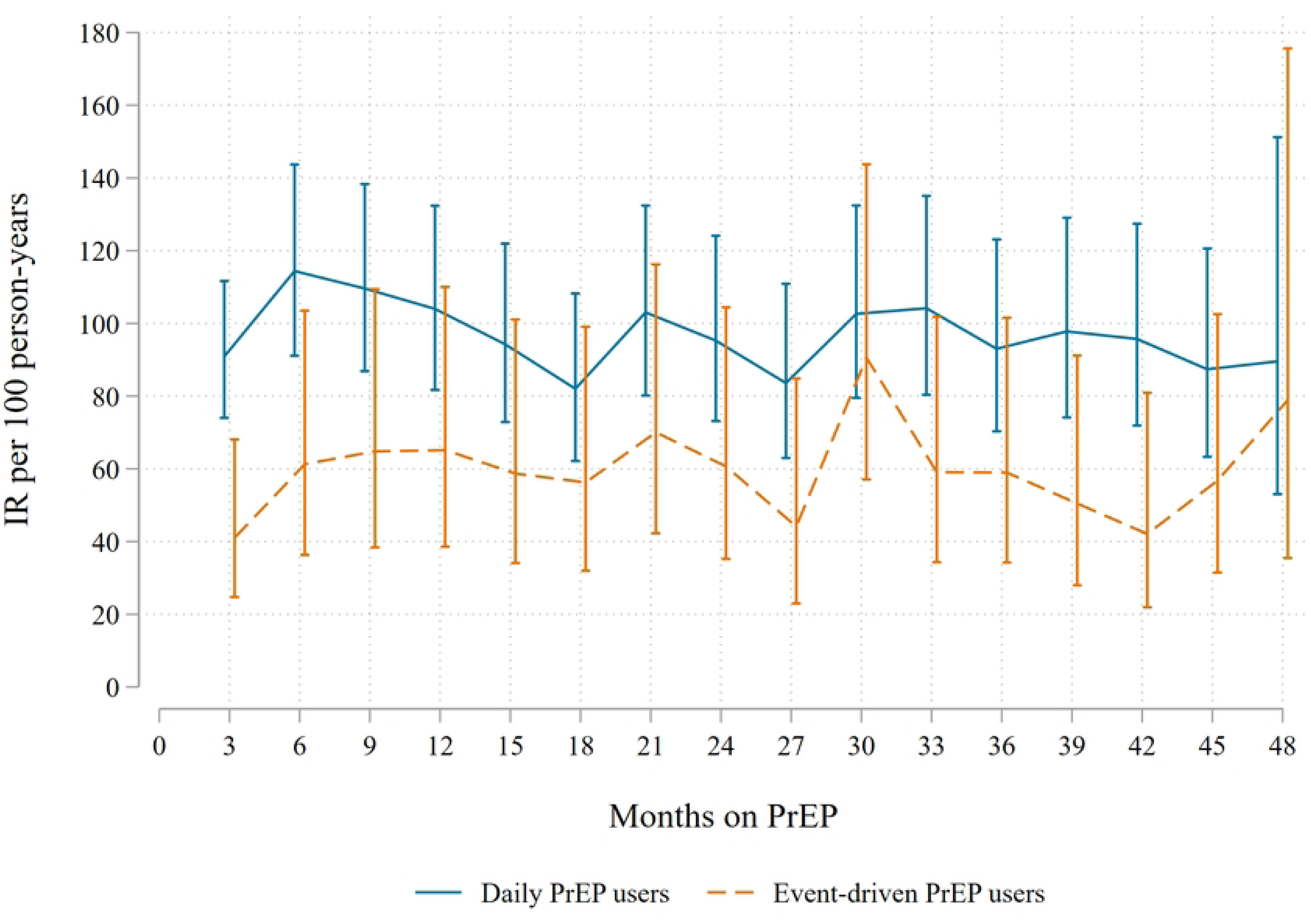
Incidence of any syphilis, gonorrhea or chlamydia among AMPrEP participants during the first four years on PrEP, AMPrEP, The Netherlands, 2015-20. Lines represent incidence rates per 100 person-years, as calculated over the previous 3 months. Vertical bars represent 95% confidence intervals. Abbreviations: AMPrEP: Amsterdam PrEP demonstration project; IR: incidence rate; PrEP: pre-exposure prophylaxis

**Table 2.** Four-year outcomes of incidence of STIs overall and by PrEP regimen, among AMPrEP participants, Amsterdam, the Netherlands, 2015-20.

|  | Total |  |  |  |  | Daily PrEP |  |  |  |  | Event-driven PrEP |  |  |  |  | p-value<sup>a</sup> |
| --- | --- | --- | --- | --- | --- | --- | --- | --- | --- | --- | --- | --- | --- | --- | --- | --- |
|  | No. of participants with ≥1 positive test | No. of positive tests | PY | IR per 100PY (95% CI) |  | No. of participants with ≥1 positive test | No. of positive tests | PY | IR per 100PY (95% CI) |  | No. of participants with ≥1 positive test | No. of positive tests | PY | IR per 100PY (95% CI) |  |  |
| **Any STI<sup>b</sup>** | 289 | 1092 | 1258 | 86.8 [81.8-92.1] |  | 246 | 891 | 914 | 97.4 [91.3-104] |  | 89 | 201 | 344 | 58.5 [50.9-67.2] |  | <0.0001* |
| **Any anal STI<sup>c</sup>** | 241 | 758 | 1258 | 60.3 [56.1-64.7] |  | 208 | 630 | 914 | 68.9 [63.7-74.5] |  | 66 | 128 | 344 | 37.2 [31.3-44.3] |  | <0.0001* |
| **Chlamydia<sup>d</sup>** | 227 | 524 | 1258 | 41.7 [38.2-45.4] |  | 197 | 432 | 914 | 47.2 [43.0-51.9] |  | 54 | 92 | 344 | 26.8 [21.8-32.8] |  | <0.0001* |
| **Gonorrhoea<sup>d</sup>** | 234 | 615 | 1258 | 48.9 [45.2-52.9] |  | 195 | 505 | 914 | 55.2 [50.6-60.3] |  | 61 | 110 | 344 | 32.0 [26.6-38.6] |  | <0.0001* |
| **Infectious syphilis<sup>e</sup>** | 111 | 140 | 1258 | 11.1 [9.4-13.1] |  | 86 | 108 | 914 | 11.8 [9.8-14.3] |  | 30 | 32 | 344 | 9.3 [6.6-13.2] |  | 0.27 |
| **HIV** | 2 | 2 | 1258 | 0.16 [0.04-0.64] |  | 2 | 2 | 914 | 0.22 [0.06-0.88] |  | 0 | 0 | 344 | 0.00 [0-1.1] |  | 0.53 |
| **HCV<sup>f,g</sup>** | 15 | 17 | 1186 | 1.4 [0.9-2.3] |  | 14 | 15 | 870 | 1.7 [1.0-2.9] |  | 2 | 2 | 315 | 0.6 [0.2-2.5] |  | 0.17 |

Abbreviations: AMPrEP: Amsterdam PrEP demonstration project; CI: confidence interval; HCV: hepatitis C virus; IR: incidence rate; LGV: lymphogranuloma venereum; PrEP: pre-exposure prophylaxis; PY: person-years; STI: sexually transmitted infection
<sup>a</sup>Two-sided p-value for crude incidence rate difference between daily and event-driven PrEP users
<sup>b</sup>Any STI: chlamydia (any location), gonorrhoea (any location), infectious syphilis (stage 1, 2 and recent latent infection)
<sup>c</sup>Any anal STI: anal chlamydia or anal gonorrhoea
<sup>d</sup>At any anatomical location
<sup>e</sup>Syphilis stage 1, stage 2 and recent latent infection
<sup>f</sup>Based on RNA positivity, and regardless of history of prior HCV infection or HCV antibody positivity
<sup>g</sup>One individual had a first HCV infection and HCV re-infection during follow-up

When stratified by regimen, similar findings were observed among daily PrEP users, but IRs were stable over yearly intervals among event-driven PrEP users (Fig 3; S5 Table). We did not observe a change in any or specific STIs over time, nor when stratified by regimen (S5 Table). Sensitivity analyses assessing STI incidence since initiating PrEP and ignoring gaps in follow-up yielded comparable results (S6 Table).

**Fig 3.**
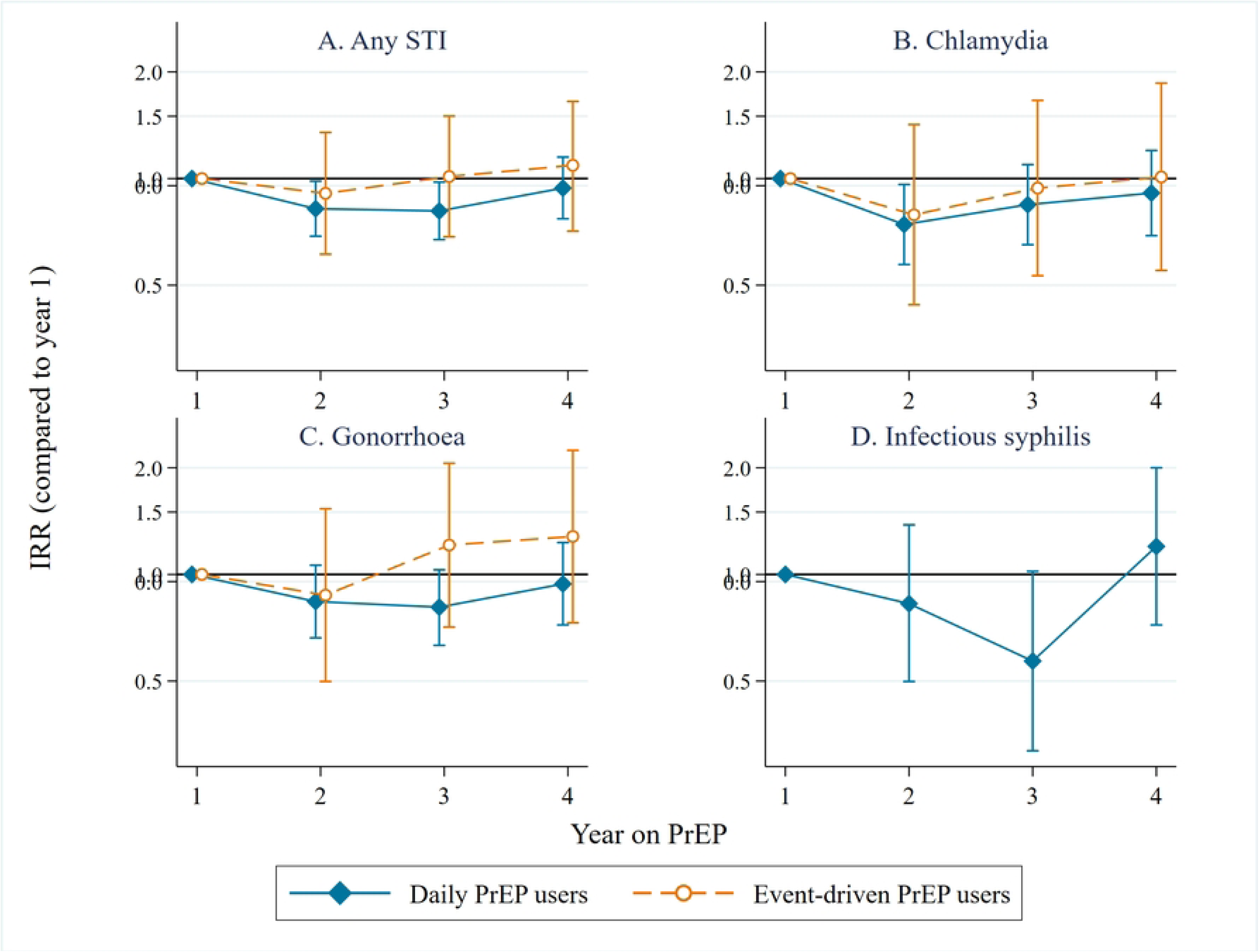
Incidence rate ratios per year on PrEP for any STI (A), chlamydia (B), gonorrhea (C) and infectious syphilis (stage 1 and 2 and recent latent; D), adjusted for age and STI testing frequency, among AMPrEP participants, Amsterdam, the Netherlands, 2015-20. Abbreviations: AMPrEP: Amsterdam PrEP demonstration project; IRR: incidence rate ratio; STI: sexually transmitted infection; NB: Year 1 is used as reference category; IRRs are adjusted for age category at baseline (i.e., <35, 35-44, ≥45 years) and individual yearly STI testing frequency. Due to a low number of newly detected infectious syphilis among event-driven PrEP users, we estimated the IRR per year on PrEP only for daily PrEP users. Vertical bars represent 95% confidence intervals.

### Incidence rates of HIV and HCV

Two daily PrEP users were diagnosed with HIV during follow-up, both in the first year since PrEP initiation, resulting in an IR of 0.16/100PY (95%CI=0.04-0.64; S4 Table). We have already reported on these cases in more detail(10, 26). In brief, one participant had discontinued PrEP several months before HIV diagnosis and the other participant was a daily PrEP user whose adherence to PrEP was confirmed to be adequate(26). No diagnosis of HIV was observed among event-driven PrEP users (IR=0/100PY, 95%CI=0-1.07; S4 Table).

17 incident HCV infections were diagnosed among 15 participants during 1186PY of follow-up (IR=1.4/100PY, 95%CI=0.89-2.30; S4 Table). Ten were primary infections and 7 reinfections. daily PrEP users accounted for 15 of these infections (IR=1.7/100PY, 95%CI=1.0-2.9) and event-driven PrEP users for two (IR=0.6/100PY, 95%CI=0.2-2.5). The number of HCV infections decreased over time: year 1, *n*=6 (IR=1.8/100PY, 95%CI=0.8-3.9); year 2, *n*=8 (IR=2.6/100PY, 95%CI=1.3-5.2); year 3, *n*=2 (IR=0.7/100PY, 95%CI=0.2-2.7); year 4, *n*=1 (IR=0.4/100PY, 95%CI=0.06-2.6).

### Regimen switching

Overall, 137/367 (37%) participants switched PrEP regimens 254 times: 141 times from daily PrEP to event-driven PrEP and 113 times from event-driven PrEP to daily PrEP use. The rate of any switch was highest in the first year (26.1/100PY) and decreased over time (IRR 0.87/year [95% CI 0.78-0.98], p=0.019) to 17.2/100PY in the fourth year (S7 Table; Fig 4).

**Fig 4:**
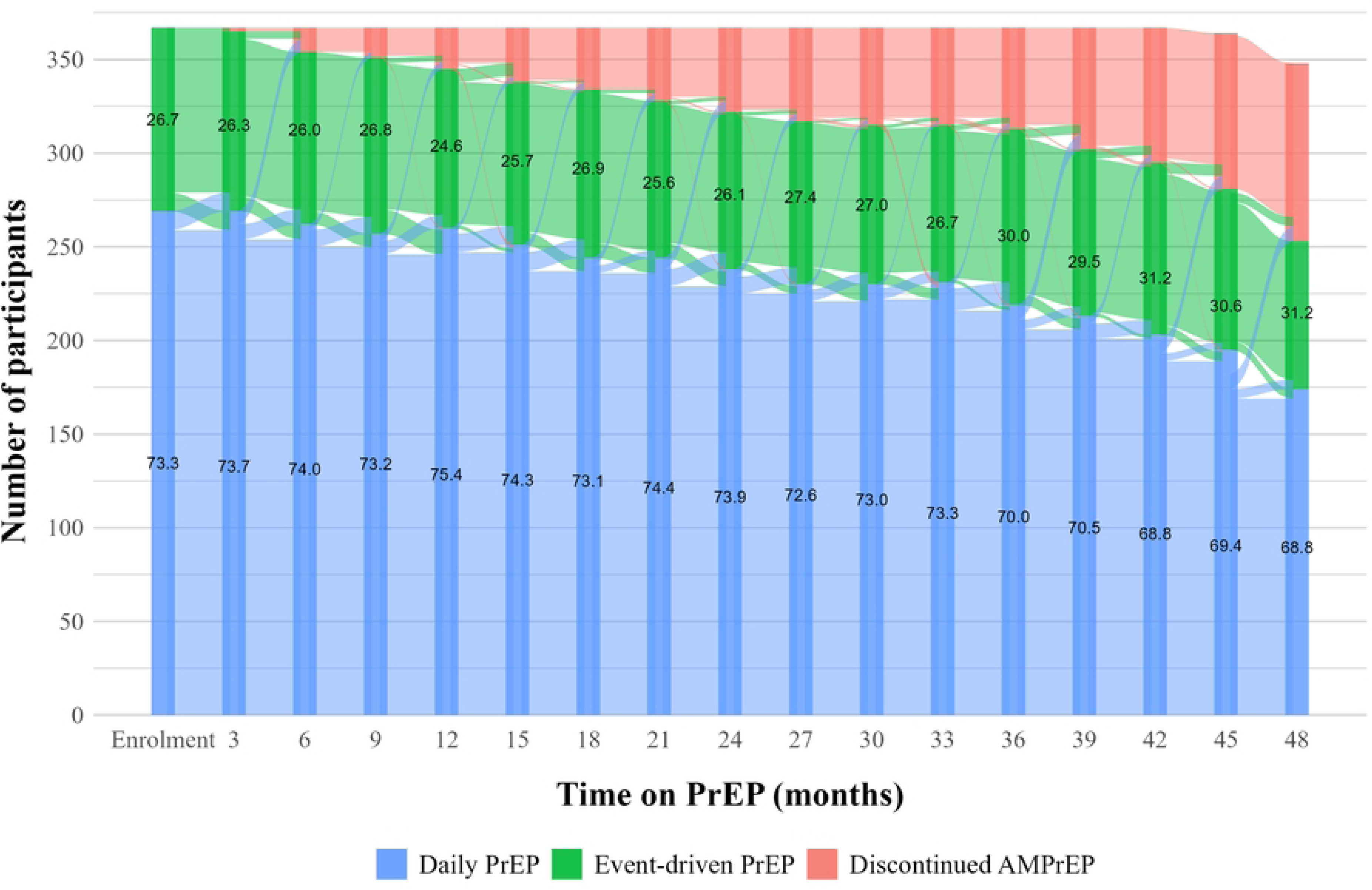
Transitions through PrEP regimens during up to four years on PrEP among AMPrEP participants, Amsterdam, the Netherlands, 2015-20. At baseline, numbers on the bars indicate the percentage of participants who intended to use daily or event-driven PrEP. At all follow-up visits, numbers on the bars indicate the percentage of participants who reported having used daily or event-driven PrEP in the past 3 months. Abbreviations: AMPrEP: Amsterdam PrEP demonstration project; PrEP: pre-exposure prophylaxis.

### Discontinuation of PrEP use

We observed 112 PrEP stops among 98/367 (27%) participants; 31 stops were subsequently followed by a restart. 43 participants had a formal study exit visit and another 43 were lost-to-follow-up. We registered 17 gaps of more than 9 months between study visits, and nine participants reported not having used PrEP for a period of at least three months, despite continuing study participation.

Median time until the first stop among those who stopped was 21 (IQR=12-35) months. Overall rate of stopping was 8.3/100PY (95%CI=6.9-10.0). In multivariable analysis, younger age (p=0.036), fewer CAS acts with casual partners (p=0.039) and not having a college-or university degree (p=0.030) were associated with earlier stopping. Being diagnosed with an STI in the past 3 months was not associated with stopping (adjusted HR=0.48, 95%CI=0.14-1.64, p=0.24; S8 Table).

### Intracellular TFV-DP concentrations

Among daily PrEP users, median TFV-DP concentration at 3 or 6 months was 1263 fmol/punch (IQR=1000-1619; n=240), at 12 months 1299 fmol/punch (IQR=1021-1627; n=259), at 24 months 1288 fmol/punch (IQR=1005-1617; n=223) and at 48 months 1693 fmol/punch (IQR=1310-2252; n=127). At 12 months, 93% (n=240/259) of participants had a TFV-DP concentration ≥700 fmol/punch, at 24 months 90% (n=200/223) and at 48 months 94% (n=120/127).

## Discussion

Over the first four years of PrEP use among participants of this prospective demonstration cohort in Amsterdam, the Netherlands, the number of CAS acts with casual partners remained stable, and the total number of sex partners decreased over time. STI incidence was high, but stable over time.

Therefore, these findings provide no indication of risk compensation in this cohort. Incidence of HIV was very low and the two incident infections occurred during the first year on PrEP. Objectively measured adherence among daily PrEP users remained well above the protective threshold during study follow-up for the large majority of participants. Retention at 48 months was high (68%). Thus, effectiveness of PrEP continues beyond the previously reported 2-year results in our and other demonstration studies(8, 10, 27). This supports implementation of low-threshold PrEP services including STI counselling, testing and treatment at regular intervals.

PrEP programme policies select for people who are behaviourally susceptible for HIV and therefore these people are prone to acquire other STIs. As AMPrEP was initiated in 2015, before the European Medicine Agency approved TDF/FTC for PrEP in July 2016, there was no other formal way to acquire PrEP in the Netherlands at the time. The lack in PrEP availability likely resulted in inclusion of a group of early PrEP adopters.

We assessed multiple sexual behaviour measures. We considered the number of CAS acts with casual partners as the most relevant factor in the context of HIV acquisition. The rate of this behaviour remained constant over four years of follow-up in both daily PrEP and event-driven PrEP users, with expectedly higher numbers among daily PrEP users. However, the total number of sex partners decreased over time, as previously observed by Molina et al. (2022)(8). Grant et al. (2010) noted a reduction of sex partners with whom participants had receptive intercourse over a median follow-up of 1.2 years, and suggested that the services around PrEP use (e.g. counselling) or taking the pill itself could serve as a reminder of HIV risk and contribute to choosing this “safer behaviour”(1). Reyniers et al. (2021) suggest a broader paradigm of improved sexual health brought about by PrEP, through empowering its users to more actively engage in their sexual health(28). AMPrEP participants had the opportunity to reflect on their sexual activity with nurses and physicians during each three-monthly consultation. This paradigm of sexual empowerment, leading to more considered sexual decisions, could explain why we observed a stable number of anal sex acts combined with fewer sex partners over time in our cohort.

AMPrEP and the Be-PrEP-ared project in Belgium were the first prospective demonstration projects to offer participants the choice between daily PrEP and event-driven PrEP, including the option to switch between regimens. We observed a high and stable incidence of STIs, similar to other early PrEP studies(29–31). We noted this especially among daily PrEP users, as well as higher numbers of sex partners and CAS acts compared to event-driven PrEP users, in agreement with an earlier, pooled analyses of AMPrEP and Be-PrEP-ared over the first 28 months(32). daily PrEP appears to be used during periods with more frequent sexual contacts or less condom use, coinciding with an increase in the chance to acquire HIV and STIs. This suggests that PrEP users are capable of deciding which regimen to use, depending on their sexual behaviour.

The low HIV incidence is likely a direct result of high PrEP adherence, as demonstrated by high median levels of TFV-DP around the level of perfect adherence. These levels were also well above the protective thresholds at all time points up to 4 years after PrEP initiation in the majority of participants. A subgroup of participants was included in a nested RCT assessing the effect of an app providing visualised feedback to increase adherence, as reported previously(23, 33), which could have possibly been a contributing factor to adherence. The absence of any HIV infection after one year on PrEP provides further evidence that PrEP use can be sustainable and efficacious in preventing HIV over the longer course. This finding is in line with other studies with less follow-up time(8, 27).

HCV prevalence was high among participants initiating PrEP in AMPrEP(12) as described in previous analyses of our cohort, but during follow-up its incidence appeared to decrease over time, parallel to the decrease seen in the Dutch population with HIV since direct-acting antivirals for HCV became widely available in 2015(34).

In our cohort, a substantial proportion of participants switched PrEP regimens once or multiple times, especially during the first year of PrEP use. Retention to PrEP remained high over four years and the vast majority of the daily PrEP users had high adherence levels at each measurement. The majority of participants that stopped using PrEP, did so because of low self-perceived need for PrEP(35). However, there can be discordance between self-perceived and actual need for PrEP(36). This can leave ex-PrEP users vulnerable to HIV, as supported by reports of high HIV incidence among people who discontinued PrEP(37), stressing the importance of low-threshold access to PrEP and adherence and persistence counselling so PrEP users may discontinue PrEP well-informed.

A major strength of this study is the long follow-up time of up to four years. In real-world settings, many are likely to use PrEP for several years and very little data were available on long term (i.e. longer than two years) PrEP use. Second, AMPrEP was a prospective, observational cohort, enabling a prospective assessment of outcome measures in which STI diagnoses made in-between study visits were included. Third, AMPrEP was among the first two demonstration projects allowing participants to choose between daily PrEP and event-driven PrEP use, adapt their PrEP use to match their need of protection, allowing an independent and long-term assessment of both regimens and inter-regimen switching behaviour.

We acknowledge some limitations. The AMPrEP cohort officially closed at 1 December 2020, but due to COVID-19 measures, we censored data after 15 March 2020, to exclude effects on sexual behaviour and STI incidence resulting from the COVID-19 pandemic. Therefore we were not able to include up to five years of follow-up nor the official end-of-study visits. Second, this cohort presumably included a high proportion of early adopters. Participants were relatively old, and mostly identified as men, white, and were college or university educated and they are unlikely to represent the broader population of MSM and transgender women who could benefit from PrEP. We were only able to include two transgender women. Future PrEP studies should aim to include a more representative sample for the population susceptible to HIV.

In conclusion, oral PrEP is a very effective HIV prevention strategy over a long period of use. We did not observe increasing numbers of sex partners, condomless anal sex acts, or STI incidence over time. These results support implementation of low-threshold PrEP services, including regular counselling, STI testing and treatment. People susceptible to HIV should be empowered to choose their preferred HIV prevention strategies, including PrEP.

## Data Availability

Data are available upon reasonable request. The AMPrEP data are owned by the Public Health Service of Amsterdam. Original data can be requested by submitting a study proposal to the steering committee of AMPrEP. The proposal format can be obtained from. Request for further information can also be submitted through the same email address. The AMPrEP steering committee verifies each proposal for compatibility with general objectives, ethical approval and informed consent forms of the AMPrEP study and potential overlap with ongoing studies. There are no restrictions to obtaining the data and all data requests will be processed in a similar way.

## Acknowledgements

We thank all AMPrEP participants, members of the advisory board and community engagement group. Additionally, we thank Roel Achterbergh, Ertan Ersan, Michelle Kroone, Dominique Loomans, Kees de Jong, Ilya Peters, Princella Felipa, Myra van Leeuwen, Jason Schouten, Kenneth Yap, Hanne Zimmermann and all members of the H-TEAM consortium (S9 Annex).

## Contributors

EH, MP and MSvdL contributed to design of the AMPrEP demonstration project and acquired funding. EH, MP, MSvdL and MvdE contributed to the study design. LC and EW conducted the data management. EW performed the statistical analyses under supervision of AB and VJ. MvdE drafted the manuscript under supervision of MSvdL. All authors contributed to the interpretation of results, critically revised the manuscript and approved the final version for publication.

## Ethics approval

The study was approved by the ethics board of the Amsterdam University Medical Centers, location AMC, Amsterdam, The Netherlands (NL49504.018.14). Participants gave written informed consent to participate in the study before taking part.

## Data sharing

Data are available upon reasonable request. The AMPrEP data are owned by the Public Health Service of Amsterdam. Original data can be requested by submitting a study proposal to the steering committee of AMPrEP. The proposal format can be obtained from.

Request for further information can also be submitted through the same email address. The AMPrEP steering committee verifies each proposal for compatibility with general objectives, ethical approval and informed consent forms of the AMPrEP study and potential overlap with ongoing studies. There are no restrictions to obtaining the data and all data requests will be processed in a similar way.

